# Predictors of glycemic worsening in the next year in adults with screen-detected type 2 diabetes

**DOI:** 10.1101/2024.04.25.24306391

**Authors:** Rebecca Schneider Aguirre, Tamara S Hannon, Robert V Considine, Yash Patel, M Sue Kirkman, Kieren J Mather

## Abstract

**Background and Aims:** Identifying simple markers of risk for worsening glucose can allow care providers to target therapeutic interventions according to risk of worsening glycemic control. We aimed to determine which routine clinical measures herald near-term glycemic worsening in early type 2 diabetes(T2D).

**Methods:** The Early Diabetes Intervention Program (EDIP) was a clinical trial in individuals with screen-detected T2D [HbA1C 6.3+0.63%(45+5mmol/mol)]. During the trial some participants experienced worsening fasting blood glucose (FBG). We investigated the time course of FBG, HbA1c, weight, and other clinical factors to determine which might herald glycemic worsening over the next year.

**Results:** Progressors (62/219, 28.5%) had higher FBG than non-progressors at baseline [118 vs 130mg/dL (6.6 vs 7.2 mmol/L), p=<0.001]. FBG was stable except in the year of progression, when progressors exhibited a large 1-year rise [mean change 14.2mg/dL(0.79 mmol/L)]. Current FBG and antecedent year change in FBG were associated with progression(p<0.01), although the magnitude of change was too small to be of clinical utility (0.19 mg/dL; 0.01 mmol/L). Current or antecedent year change in HbA1c, weight, TG or HDL were not associated with progression. In the year of glycemic worsening, rising glucose was strongly associated with a concurrent increase in weight (p<0.001).

**Conclusions:** Elevated FBG but not HbA1c identified individuals at risk for imminent glycemic worsening; the subsequent large rise in glucose was associated with a short-term increase in weight. Glucose and weight surveillance provide actionable information for those caring for patients with early diabetes.

**Highlights:**

- Recent change in fasting blood sugar is associated with near-term worsening of glycemia
- Changes in weight, insulin, HbA1C were not informative for anticipating near-term worsening of glycemia
- Increasing glucose was strongly associated with concurrent weight gain

## 1.1 Introduction

The progressive dysglycemia underlying the development of Type 2 diabetes (T2D) is characterized by a sequence starting with normoglycemic insulin resistance, followed by impaired fasting glucose (IFG) and/or impaired glucose tolerance (IGT) and then finally a transition to overt diabetes (1). Among obese individuals with pre-diabetes, approximately 5-10% will progress to diabetes yearly, with a cumulative risk of progression of up to 70% (2). A similar process of progressive loss of glycemic control is also seen in established diabetes, beginning from the earliest stages of diabetes (1, 3). Within groups with pre-diabetes, as well as those with established diabetes, individuals exhibit varying rates of glycemic worsening. Epidemiologic factors do not appear to be sufficient to identify those at greatest risk of near-term progression (4). Clinically accessible tools are needed to identify these individuals, in order to intervene effectively.

The current knowledge of epidemiologic predictors of glycemic worsening comes from natural history studies. Prominent among these, the longitudinal Whitehall II cohort study screened individuals for diabetes and then followed those without diabetes for up to 13 years (5). While baseline glycemia differed significantly between non-progressors and progressors to diabetes, these differences in baseline average FBG values were less than 10 mg/dl (0.56 mM) between the two groups, a difference that is easily overlooked in clinical surveillance. The near-term changes in FBG were more dramatically different between groups, suggesting that prospective monitoring of glycemia over time could be informative for prediction of progression.

The Early Diabetes Intervention Program (EDIP) was a double-blind placebo-controlled intervention trial that identified individuals with screen-detected diabetes (fasting glucose <140 mg/dl or <7.8 mM, 2 hr OGTT glucose >200 mg/dL or >11.1 mM) according to diagnostic thresholds at time of study design (1997), and evaluated whether acarbose could delay the rise of fasting glucose compared to placebo. In the primary analysis, no statistical difference was found between acarbose and placebo in the rate of glycemic worsening over 5 years (6) and there were no improvements in β-cell function with acarbose treatment (7). At enrollment, the average fasting glucose was 121±14 mg/dL (6.7 +/-0.78 mM) and the HbA1c was 6.3+0.63% (45 + 5.0 mmol/mol); although these results clearly identify dysglycemia, very few of these individuals would be diagnosed with diabetes in usual care. This study population is informative for questions relating to worsening glycemia at the threshold between prediabetes and diabetes.

The goal of the current analyses was to evaluate possible predictors of worsening glycemia that are easily accessible to those treating the majority of patients with prediabetes and diabetes, namely primary care providers. We evaluated measures that could be used to identify imminent worsening of glycemia, including current values and 1-year antecedent changes in, glucose, HbA1c, weight and the TG/HDL ratio. Previous EDIP analyses used Cox proportional hazards time-to-event analyses to identify risk factors associated with study-defined progression of fasting blood glucose at the population level (7). Here we have focused on measures antecedent to the progression event, in order to isolate factors that identify risk of near-term progression that might be used in prospective monitoring to identify individuals at risk of near-term worsening. Our hypothesis was that both the absolute levels as well as the recent change in these parameters would be predictive of glycemic worsening over the upcoming year.

## 2.1 Methods

EDIP was undertaken at two study sites, Indiana University School of Medicine and Washington University School of Medicine. The study was approved by the institutional review boards at both sites, and all subjects provided written informed consent for their participation. Inclusion and exclusion criteria, and general methods for the EDIP trial have been previously published (6). Briefly, participants were recruited from asymptomatic individuals without known diabetes, who were screened with a 2-hour 75g oral glucose tolerance test (OGTT). Diabetes was defined with a 2hr blood glucose value >200 mg/dL or >11.1 mM, and for study purposes participants were required to have a fasting glucose value below 140 mg/dL (7.8 mM). Following randomization to either acarbose or placebo, subjects were followed with quarterly measurements of fasting glucose to ascertain glycemic progression, defined as two consecutive quarterly FBG values above 140 mg/dL. If subjects met the primary endpoint, their participation was terminated (6).

### 2.1.1. Calculations

Fasting insulin values were the average of the two baseline values measured at the time of OGTT procedures. Insulin sensitivity was estimated using inverse fasting insulin, combined with concurrent glucose readings into a Homeostasis Model Assessment of Insulin Resistance (HOMA-IR) calculation or combined with the insulinogenic index (IGI=(ins30-ins0)/(gluc30-gluc0)) from the OGTT to derive an oral disposition index (oDI = IGI * HOMA-I(8-10)⍰. Where this calculation produced negative values (a numerically possible but physiologically uninterpretable result), those were set to missing.

### 2.1.2 Assays

Glucose concentrations were determined using a glucose oxidase method (YSI, Yellow Springs, OH). HbA1c was measured using an immunoturbidometric assay (Roche Diagnostics, Indianapolis, IN). Total and HDL cholesterol and triglycerides were measured using an enzymatic end-point assay (Roche Diagnostics). Insulin and proinsulin were measured by radioimmunoassay (Linco Research). All laboratory assays, other than the on-site YSI plasma glucose measurements, were done at the central study laboratory at the Indiana University School of Medicine.

### 2.1.3 Statistical analysis

Demographic and metabolic parameters were compared between groups using independent t-tests. Baseline characteristics used study-entry data points; other data points evaluated realigned data as described below.

To allow for predictive analysis, time zero was defined as the time of censoring, representing either the time of progression or the last study visit with glucose measurements. Data were recorded such that the timing of antecedent measurements was expressed relative to the time of censoring (i.e. censoring minus 1 year, censoring minus 2 years, etc). Censoring events could happen on quarter-year intervals; where these did not fall on a study year timepoint, for purposes of alignment with annually collected data these were counted as their nearest subsequent study year. Changes over one year were calculated by taking each individual’s change from the temporally distant measure to the next more proximal measure, e.g. Year −2 minus Year −1, so that values represent prospective changes forward over time. We have annotated this so that each year interval is represented by Δa,b, representing the interval change between “a” as the beginning time point and “b” as the ending time point in the interval. For example, Δ-2,-1 represents the change from two years prior to censor to one year prior to censor.

Logistic regression was used to evaluate factors associated with subsequent progression events.

We focused on evaluating measures that are readily available in primary care clinics, namely weight, fasting glucose, HbA1C, insulin concentrations, and TG/HDL ratios. We also evaluated simple measures of insulin sensitivity (HOMA-IR) and β-cell function (IGI and oDI), recognizing that these are not generally performed in primary care clinics. We focused on the potential impact of ‘current’ measures in a given time frame prior to progression, and on 1-year changes prior to this time point, on the basis that these would be realistically available in usual clinical practice to identify individuals at risk of progression.

Linear mixed modeling was performed to evaluate potential determinants of the change in fasting blood glucose in the year of progression, with the rationale that such factors could be a focus of clinical intervention.

The data were primarily analyzed using the entire study population. We also performed parallel analyses in the subset of the population that could be evaluated using progression above 126 mg/dL (7.0 mmol/L), as this is the current diagnostic threshold between impaired fasting glucose and diabetes. To accomplish this, we selected the subset of participants with FBG < 126 mg/dl at baseline (<7.0 mmol/L), and defined progression fasting blood glucose values >126 mg/dL (>7.0 mmol/L). Analyses were repeated as detailed above.

SPSS software was used to perform all statistical analysis (Version 24, IBM Chicago IL). Two-sided *p* values <0.05 were considered statistically significant.

## 3.1 Results

### 3.1.1. Baseline Characteristics

Characteristics of study participants have been previously published (6). **Table 1** summarizes the relevant baseline characteristics, comparing those who met criteria for progression [i.e. exhibited fasting glucose >140 mg/dl (>7.8 mmol/l) on 2 consecutive quarterly assessments], versus those who did not, in the 5-year observation period. Baseline demographics, weight, and BMI were not different between the non-progressors and progressors. Notably, the two groups also did not differ at study entry with respect to baseline HbA1C levels, 2-hour oral glucose tolerance test results, fasting insulin levels, HOMA-IR (insulin sensitivity) and Oral Disposition Index (β-cell function). Fasting blood glucose (FBG) concentration at study entry was the only statistically different baseline parameter between non-progressors and progressors [118.4 mg/dl (6.6 mmol/l) vs 129.5 mg/dl (7.2 mmol/l), respectively; p<0.0001]. EDIP was conceived when the criteria for diagnosis of diabetes included a FBG >140 mg/dL (>7.8 mmol/l); we also analyzed the baseline characteristics of participants excluding those who had a FBG of >126 mg/dL (>7.0 mmol/l) at study entry, with similar results overall (data not shown).

**Table 1:** Baseline demographic and metabolic characteristics of the population. Summary values are presented as mean±SD. The p-value is comparing the non-progressors and progressors (2-tailed, equal variances assumed). Abbreviations: Af-Amer, African-American; BMI, body mass index; Cauc, Caucasian; DI, disposition index; FBG, fasting blood glucose; HDL, high density lipoprotein; HbA1C, hemoglobin A1C; HOMA-IR, homeostatic model assessment-insulin resistance; Ins, insulin; oDI, oral disposition index; OGTT-2h, oral glucose tolerance test – 2-hour blood glucose; TG, triglycerides.

| Baseline Characteristic | All (n = 219) | Non-progressors (n = 157) | Progressors (n = 62) | p-value ($\chi^2$ analysis or t-test) |
| --- | --- | --- | --- | --- |
| Age (years,) | 53.7±11.4 | 54.1 ± 11.3 | 52.6 ± 11.5 | 0.36 |
| Sex:<br>Male/Female | 74/145 | 48 / 109 | 26/36 | 0.12 |
| Race:<br>Cauc/Af-Am/Other | 158/53/9 | 117/42/9 | 51/11/0 | 0.25 |
| Weight (kg,) | 98.7±21.2 | 97.3 ± 21.0 | 102.4 ± 21.2 | 0.11 |
| BMI (kg/m<sup>2</sup>) | 35.2±7.1 | 35.0 ± 7.2 | 35.7 ± 7.0 | 0.52 |
| FBG (mg/dL) | 121.5±13.6 | 118.4 ± 13.9 | 129.5 ± 9.2 | <0.00001 |
| FBG (mmol/l) | 6.7 ± 0.8 | 6.6 ± 0.8 | 7.2 ± 0.5 | <0.00001 |
| HbA1C (%) | 6.3±0.63% | 6.3 ± 0.65% | 6.4 ± 0.57% | 0.56 |
| HbA1c (mmol/mol) | 45±5 | 45±5 | 45±4 | 0.56 |
| OGTT-2h (mg/dl) | 236.2±31.1 | 235.0 ± 28.7 | 239.3 ± 36.5 | 0.37 |
| OGTT-2h (mmol/l) | 13.1 ± 1.7 | 13.0 ± 1.6 | 13.2 ± 2.0 | 0.37 |
| Fasting Ins (mU/L) | 20.3±13.5 | 19.9 ± 13.7 | 21.2 ± 13.0 | 0.52 |
| Fasting Ins (pmol/l) | 141.0 ± 93.8 | 138.2 ± 95.1 | 147.2 ± 90.3 | 0.52 |
| HOMA-IR | 6.1 ± 4.2 | 5.9 ± 4.3 | 6.7 ± 3.9 | 0.19 |
| oDI | 4.2±9.2 | 4.7 ± 8.9 | 2.9 ± 9.6 | 0.20 |
| TG (mg/dl) | 193.7 ± 102.3 | 186.4 ± 96.6 | 212.4 ± 114.6 | 0.10 |
| TG (mmol/L) | 2.2 ± 1.2 | 2.1 ± 1.1 | 2.4 ± 1.3 | 0.10 |
| HDL (mg/dl) | 38.1 ± 8.9 | 38.7 ± 8.5 | 36.4 ± 9.9 | 0.09 |
| HDL (mmol/L) | (1.0 ± 0.2) | (1.0 ± 0.2) | (1.0 ± 0.3) | 0.09 |

### 3.1.2. Progressors are defined by a large change in fasting blood glucose

Figure 1. presents the average FBG, HbA1c and weight from 4 years prior to censor. A few features are of note. First, the differences between progressors and non-progressors in the overall mean fasting glucose values was modest, with elevated values in both groups that were relatively stable over time until the year of progression. Second, the rise of fasting glucose that defines the progression event among progressors was not a modest change; it was instead a large change that negates the notion that progressors were simply closer to the threshold. Third, while HbA1c values rose overall in both groups, this was not a feature that distinguished these groups. Fourth, the time course of weight change was not the same between groups, although again a general pattern of increase over time was seen in both groups.

**Figure 1.**
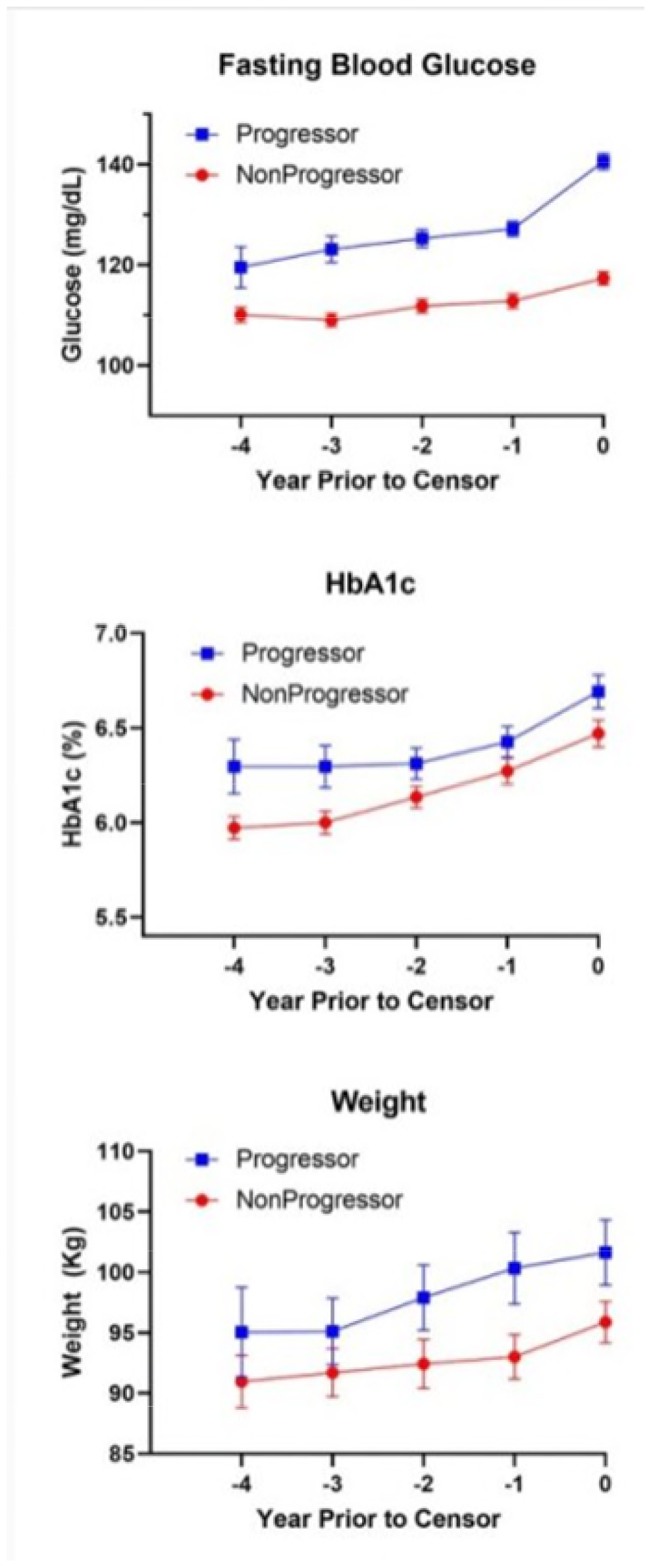
Time course of Fasting Blood Glucose, HbA1c and Weight in the years preceding progression.

### 3.1.3. Fasting blood sugar, but not A1C, is most predictive of impending progression

We used logistic regression analysis to evaluate whether various clinical measures could predict progression (**Table 2)**. In univariate analyses, FBG (Model 1) measured prior to the progression event predicted progression; adding one-year antecedent changes improved the model (Model 2). It is notable that HbA1C was not predictive of glycemic worsening, either in isolation (data not shown) or as part of a model (Models 3, 4). In fact, substituting HbA1C (Models 3, 4) for FBG (Models 1, 2) rendered the models statistically non-significant. Although the ΔFBG for the Δ-2,-1 interval (beginning 2 years prior to progression) was significant, the slope was unexpectedly inverse and the absolute change in that year was very small on average (0.19 mg/dl (0.01 mmol/l); **Figure 1**), suggesting that the directionality estimate is unstable. Regardless, such changes are unlikely detectable in routine clinical surveillance of an individual. Weight and lipid concentrations, and their antecedent changes, did not provide independent additional information about risk of progression. Models 3 and 4 substitute HbA1c for the fasting glucose readings; in both models this alternative measure of glucose control did not achieve significance, and in fact the models themselves no longer reached significance. On balance these analyses point to informative data from the fasting blood glucose but not HbA1c in the year prior to progression. Weight and lipid readings in the same interval did not augment the information provided by fasting glucose.

**Table 2:** Logistic regression analysis evaluating predictors of progression the year prior to progression events. 0, time zero or censor; −1, one year prior to sensor; −2, two years prior to censor; −3, three years prior to censor; Δ0,-1, change over year ending in censor; Δ-1,-2, change over one year ending in the year prior to censor. Model 1, Year −1 determinants. Model 2; Year −1 and Δ0,-1 determinants. Models 3 and 4 are the same as Models 1 and 2 respectively, but using HbA1c instead of FBG. FBG, fasting blood glucose; HbA1c, hemoglobin A1c; HDL, high-density lipoprotein; TG, triglycerides.

| Model # | p model | R<sup>2</sup> | Variables included | Standardized coefficient b [SE(b)] | p coefficient |
| --- | --- | --- | --- | --- | --- |
| **1** | <0.0001 | 0.287 | Weight (-1) | 0.005 [0.011] | 0.677 |
|  |  |  | FBG (-1) | 0.078 [0.016] | <0.0001 |
|  |  |  | HDL (-1) | 0.008 [0.020] | 0.681 |
|  |  |  | TG (-1) | 0.002 [0.002] | 0.219 |
| **2** | <0.0001 | 0.385 | Weight (-1) | -0.003 [0.014] | 0.855 |
|  |  |  | FBG (-1) | 0.125 [0.028] | <0.0001 |
|  |  |  | HDL (-1) | 0.009 [0.026] | 0.6726 |
|  |  |  | TG (-1) | 0.004 [0.003] | 0.284 |
| | | | ΔWeight ($\Delta -2$, -1) | 0.049 [0.079] | 0.534 |
| | | | ΔFBG ($\Delta -2$, -1) | -0.086 [0.030] | 0.004 |
| | | | ΔHDL ($\Delta -2$, -1) | 0.049 [0.007] | 0.176 |
| | | | ΔTG ($\Delta -2$, -1) | -0.003 [0.004] | 0.444 |
| **3** | 0.095 | 0.067 | Weight (-1) | 0.015 [0.009] | 0.102 |
|  |  |  | HbA1c (-1) | 0.301 [0.249] | 0.227 |
|  |  |  | HDL (-1) | 0.001 [0.018] | 0.956 |
|  |  |  | TG (-1) | 0.003 [0.002] | 0.113 |
| **4** | 0.263 | 0.120 | Weight (-1) | 0.012 [0.012] | 0.296 |
|  |  |  | HbA1c (-1) | 0.847 [0.459] | 0.065 |
|  |  |  | HDL (-1) | 0.009 [0.024] | 0.719 |
|  |  |  | TG (-1) | 0.004 [0.003] | 0.159 |
| | | | ΔWeight ($\Delta -2$, -1) | -0.026 [0.057] | 0.663 |
| | | | ΔHbA1C ($\Delta -2$, -1) | -0.786 [0.526] | 0.135 |
| | | | ΔHDL ($\Delta -2$, -1) | 0.053 [0.034] | 0.112 |
| | | | ΔTG ($\Delta -2$, -1) | -0.002 [0.003] | 0.474 |

### 3.1.4. Increase in glucose is associated with increase in weight

Progression above a pre-defined FBG threshold could result from modest increases in FBG in those with near-threshold values (’creepers’), or from a more significant increment in FBG in those further from the threshold (’leapers’). Data from **Table 2** and **Figure 1** suggest that the most prevalent mode of progression involved large change. We examined if there were parameters at the Year −1 timepoint that were associated with a >5 mg/dl (>0.3 mmol/l) or >10 mg/dl (>0.6 mmol/l) change in FBG. Between those who had this change and those who did not, there were no significant differences in 2h OGTT glucose, HbA1C, HDL, TGs, weight, or insulin concentrations (data not shown). Linear regression using either a >5 mg/dl (>0.3 mmol/l) or >10 mg/dl (>0.6 mmol/l) change in FBG as the dependent variable similarly failed to identify any of the evaluated variables as significantly related to these thresholds of change (data not shown).

We next sought identify factors associated with the change in FBG in the year of progression (i.e. concurrent changes). We used linear mixed model analyses to examine contributions of changes in weight, fasting insulin levels, and β cell function (oDI), including testing whether these factors were different in progressors versus non-progressors. **Table 3** shows that change in weight was significantly and directly associated with the change in fasting glucose, and that this effect differed between progressors and non-progressors. The changes in fasting insulin and β-cell function were not related to the increase in glucose either individually or when the interaction with progression group was evaluated. These analyses highlight increases in weight as the major phenomenon associated with increases in glucose at the time of glycemic progression.

**Table 3.** Mixed Model evaluating changes associated with increase in fasting glucose during the year of progression. Δ indicates the change in the year of progression. HDL, high-density lipoprotein; oDI, oral disposition index (a measure of insulin secretion).

| Variables included | Estimate | Standard Error | Significance |
| --- | --- | --- | --- |
| ΔWeight | 2.714 | 0.626 | <0.001 |
| ΔInsulin | 0.076 | 0.143 | 0.796 |
| ΔoDI | 0.281 | 0.354 | 0.166 |
| Progressor x ΔWeight | -2.171 | 0.731 | <0.01 |
| Progressor x ΔIns | -0.104 | 0.187 | 0.578 |
| Progressor x ΔoDI | 0.014 | 0.411 | 0.973 |

## 4.1 Discussion

The current analyses aimed to identify predictors of imminent glycemic worsening in individuals with early diabetes, characterized by glycemic measures near the diagnostic threshold for diabetes.

Progressors had higher fasting glucose and higher HbA1c than non-progressors, with an important and clinically recognizable difference between values in these groups. The progression events occurred within a short time frame; no antecedent change in glucose or HbA1c was seen that could be used to more proactively identify imminent glycemic worsening. No association with ‘current’ weight or TG/HDL ratio, or antecedent change in weight or TG/HDL ratio, other commonly assessed clinical measures, was seen. Notably, our analyses showed that the change in fasting glucose during the year of progression was strongly associated with concurrent change in weight. This is a clinically useful observation, in that it identifies a target for intervention that can be applied in efforts to prevent glycemic worsening among those with risk levels of glycemia.

These observations are parallel to those of the Whitehall II study, which followed a European population with pre-diabetes for progression to diabetes and found in a similar reverse-time analysis that progressors had modestly higher glucose values at baseline that were overall stable in the years before progression, and then experienced progression as a short-term event (5). The observation of elevation of glycemia as a dominant predictor of risk of glycemic worsening is in line with most epidemiologic and prospective studies (11-13). In such studies, weight, insulin resistance, and/or measures of dyslipidemia within the construct of the metabolic syndrome are generally also associated with risk of glycemic progression. The current study population more cleanly isolates general epidemiologic risks from factors that predict near-term change, and in this way better focuses the clinician on factors of immediate importance.

Even though current weight and recent change in weight did not identify risk of near-term progression, the gain in weight in the year of progression was strongly associated with the worsening in glycemia that defined the progression event. Also, greater weight gain had occurred among the progressors in the antecedent 2 years than in the non-progressors. This provides a key clinical message, namely that weight maintenance in diabetes is a relevant goal and target for intervention. The strong diabetes reversal effects of intensive weight loss through diet and exercise, or through surgical interventions, are well known (14, 15) but our data suggest that the corollary clinical goal of weight maintenance and avoidance of weight gain also has clear value.

Importantly, HbA1C was not a significant predictor of imminent glycemic worsening. This is in part because the between-group differences were modest, and in part because HbA1c values were rising modestly in parallel in both groups (**Figure 1**). All study participants had elevated BGs after meals, as evidenced by the failed OGTT required for study entry. However, not all individuals had FBGs at the threshold for diabetes diagnosis (including current criteria). These observations highlight the differential contribution of fasting and post-prandial glucose levels to the time-averaging effect of HbA1c. The main implication for the current evaluation is that different time courses of fasting and post-prandial glucose in the time course of disease worsening require separate monitoring strategies. Therefore, clinicians should consider following FBG as well as A1C for risk stratification and targeted interventions aimed at preventing worsening dysglycemia.

These observations have simple and clear clinical implications. First, even modestly elevated glucose within the range seen in prediabetes and early diabetes puts individuals at risk for progressive worsening of glycemia. In progressors and non-progressors, measures of glycemia were generally stable over time, but progressors experienced relatively short-term progression events that resulted in a significant worsening of glycemia. These progression events were strongly associated with weight gain. Together these observations argue for routine surveillance of glycemia in prediabetes and early diabetes, with close attention to change in FBG (and not just HbA1c), paired with prospective counseling and interventions to avoid weight gain.

The principal strength of this study is the close surveillance of glycemia and other clinical factors that allowed a detailed assessment of the factors associated with near-term progression. There are a number of limitations. These include a somewhat unrepresentative, primarily Caucasian, study population. The data arise from a research trial, which entailed lifestyle counseling and regular study visits, so there may be a selection bias of the participants and potential differences in the time course of changes in this population compared to a more general patient population. The EDIP study was also conceived in 1997 when the FBG threshold for diabetes was 140 mg/dl (7.8 mmol/l), and thus the study population is different than what it would be using current diagnostic criteria. We addressed this by analyzing a subset tailored to the current diagnostic criteria (FBG <126 mg/dl or 7 mmol/l).

Reassuringly, there were no differences in the results observed in this subset compared to the main results presented.

This study extends prior observations underscoring the importance of surveillance of glucose in pre-diabetes, demonstrating that such surveillance can help identify individuals with early diabetes at risk of worsening. This study also adds to previously demonstrated findings that weight loss confers glycemic and overall health benefits in pre-diabetes and diabetes; our observations suggest that avoidance of weight gain is an equally important goal.

## 5.1 Conclusions

FBG was more informative for impending glycemic worsening than HbA1C alone, indicating that FBG may be more sensitive to use as a tool to identify risk of glycemic worsening. The effects of weight loss on risk of glycemic progression are well-known; we show here the corollary, that prevention of weight gain is a therapeutic target for halting worsening dysglycemia in those with prediabetes or early T2D as defined in usual care.

## Data Availability

All data produced in the present study are available upon reasonable request to the authors

## Abbreviations

BMI: body mass index;
EDIP: Early Diabetes Intervention Program;
FBG: fasting blood glucose;
HOMA-IR: Homeostasis Model Assessment of Insulin Resistance;
IFG: impaired fasting glucose;
IGT: impaired glucose tolerance;
IGI: insulinogenic index;
oDI: oral disposition index;
OGTT: oral glucose tolerance test;
T2D: type 2 diabetes

